# Diagnosing Community-Acquired Pneumonia: diagnostic accuracy study of a cough-centred algorithm for use in primary and acute-care consultations

**DOI:** 10.1101/2020.09.11.20190967

**Authors:** Paul Porter, Joanna Brisbane, Udantha Abeyratne, Natasha Bear, Javan Wood, Vesa Peltonen, Phillip Della, Claire Smith, Scott Claxton

**Affiliations:** School of Nursing, Midwifery and Paramedicine, Curtin University, Bentley, Western Australia, 6102; Joondalup Health Campus, Joondalup, Western Australia, 6027; School of Information Technology and Electrical Engineering, University of Queensland, Brisbane, Queensland, Australia, 4072; Institute of Health Research, University of Notre Dame, Western Australia, 6959; ResApp Health, Brisbane, Queensland, Australia, 4000; Genesis Care Sleep and Respiratory, Perth, Western Australia, 6027

**Keywords:** Community-acquired pneumonia, Diagnostic algorithm, Primary Care, Digital health

## Abstract

**Background:** Community-acquired pneumonia (CAP) is an essential consideration in patients presenting to primary care with respiratory symptoms; however, accurate diagnosis is difficult when clinical and radiologic examinations are not possible, such as during telehealth consultations.

**Aim:** To develop and test a smartphone-based algorithm for diagnosing CAP without need for clinical examination or radiology inputs.

**Design and Setting:** A prospective cohort study using data from subjects aged over 12 years presenting with acute respiratory symptoms to a hospital in Western Australia.

**Method:** Five cough audio-segments were recorded and four patient-reported symptoms (fever, acute cough, productive cough, age) were analysed by the smartphone-based algorithm to generate an immediate diagnostic output for CAP. We recruited independent cohorts to train and test the accuracy of the algorithm.

Diagnostic agreement was calculated against the confirmed discharge diagnosis of CAP by specialist physicians. Specialist radiologists reported medical imaging.

**Results:** The algorithm had high percent agreement (PA) with the clinical diagnosis of CAP in the total cohort (n=322, Positive PA=86%, Negative PA=86%, AUC=0.95); in subjects 22-65 years (n=192, PPA=86%, NPA=87%, AUC=0.94) and in subjects >65 years (n=86, PPA=86%, NPA=87.5%, AUC=0.94). Agreement was preserved across CAP severity: 85% (80/94) of subjects with CRB-65 scores 1-2, and 87% (57/65) with a score of 0, were correctly diagnosed by the algorithm.

**Conclusion:** The algorithm provides rapid and accurate diagnosis of CAP. It offers improved accuracy over current protocols when clinical evaluation is difficult. It provides increased capabilities for primary and acute care, including telehealth services, required during the COVID-19 pandemic.

**How this fits in?:** Diagnosis of community-acquired pneumonia (CAP) in the primary care setting relies upon the identification of clinical features or abnormal vital signs during a clinical examination. We have developed a smartphone-based algorithm which removes the requirement for in-person consultation and provides high-diagnostic agreement with specialist diagnosis of CAP. The algorithm requires the input of five cough-sound segments and four patient-reported symptoms and provides a result in less than one minute. With increasing momentum towards digital-first care under the NHS, tools such as this which allow remote deployment are likely to find increased merit.

## INTRODUCTION

Although community-acquired pneumonia (CAP) remains a leading cause of morbidity and mortality, an accurate diagnosis can be difficult; and is reliant on excellent clinical skills with or without radiology. Diagnosis is more difficult in the elderly where symptoms and signs may be minimal or the presentation modified by underlying comorbidities (1). Moreover, as current guidelines recommend moving towards digital consultations during the COVID-19 pandemic, diagnosing CAP is even more challenging when doctors are unable to conduct clinical examinations. There is a need to develop accurate methods to assist physicians to diagnose CAP without relying on clinical examination or radiology, with desirable features including ease of use and point-of-care results.

Globally, pneumonia is the most common cause of infectious mortality, with 2 million adults dying from lower respiratory infections in 2015 (2). CAP is an essential consideration in patients presenting with respiratory symptoms. In the UK, 5-12% of patients presenting to primary care with respiratory conditions have CAP (3). In the United States, 80% of CAP are managed as outpatients; and over 8 million CAP patients are admitted to hospital annually, with an overall mortality rate of 8.8% (4).

The diagnosis of CAP relies on the presence of select clinical features (cough, fever, sputum production, pleuritic chest pain) and abnormal vital signs (temperature, breathing and heart rates), supported by finding new infiltrates on chest x-rays (CXRs). Despite this, clinical symptoms and signs in isolation have generally performed poorly as diagnostic criteria (5). Auscultation findings alone have poor sensitivity and demonstrate poor agreement between clinicians (6-8). Vital sign measurements are recommended to rule out CAP: a systemic review in 2019 showed validity using a combination of breathing rate, heart rate and fever (-ve LR, 0.24 (0.17 to 0.34) (9).

The utility of CXRs in diagnosing CAP is questionable. Agreement on CXR interpretation between Emergency Department clinicians or GPs with radiologists is poor. CXRs have been shown to lack precision, reliability and consistency; and are not useful in determining disease aetiology (10-12). More information can be obtained from CT and ultrasound examinations, but they are costly and not readily available. In primary care settings, access to radiology may be limited and take time thus delaying the initiation of treatment.

Guidelines for the role of CXR in CAP diagnoses are inconsistent. The American Thoracic Society recommends routine CXRs for suspected CAP (13). The British Thoracic Society recommends CXRs for hospitalised patients but not for community treatment unless there are additional concerns such as inadequate response to treatment (14). The European Respiratory Society (ERS) recommends separating “definite” from “suspected” CAP based on the presence or absence of an abnormal CXR; then treating both groups with empirical antibiotics (15). Most CAPs in the UK are thus diagnosed using clinical features alone.

In 2015 we commenced a digital diagnostic program to develop algorithms that combine a mathematical analysis of cough-associated audio segments with patient-reported symptoms to identify respiratory diseases in children and adults (16-18). The forced expiratory air column produced during a cough supports a higher bandwidth than that across the chest wall which is relied upon in traditional auscultation. Sounds generated inside the lungs propagate through this air column and the pathophysiological changes caused by different respiratory conditions modulate the sound quality. The identification of unique sound signatures characteristic of different conditions, led to the development of algorithms for the diagnosis of each respiratory condition. The algorithms do not rely on the input of vital signs, clinical examination or radiology.

Pilot studies using this method demonstrated a sensitivity of 94% and specificity of 88% for differentiating pneumonia from no disease. Subsequently, we published the accuracy of an algorithm to differentiate paediatric patients with croup, asthma, pneumonia, bronchiolitis and upper/lower respiratory disease (16). The PPA and NPA for pneumonia in children aged 2-12yrs were 85% and 80% and for 28 days-2yrs, 100% and 97%. The aim of the current study was to develop and test the performance of an algorithm to diagnose CAP in an adolescent and adult population presenting to acute-care settings with respiratory symptoms.

## METHODS

### Study design

A prospective study comparing the diagnostic accuracy of an automated algorithm to clinical diagnosis of CAP in subjects >12 years attending acute-care units in an Australian hospital.

### Study Population and Setting

Between January 2016 and March 2019, we recruited two cohorts as convenience samples from a large, general hospital in Western Australia. Data from the first cohort was used to develop and optimise the software algorithm. We used the second, independent, cohort to test the optimised algorithm.

Subjects came from multiple hospital departments; including the primary-care unit, emergency department, and inpatient wards. Subjects were approached if they presented with signs or symptoms of acute respiratory disease. All subjects who met the inclusion criteria were eligible, except for those with documented COPD or restrictive lung disease (we have previously developed diagnostic algorithms for these). Inclusion/exclusion criteria are presented in Table 1.

**Table 1.** Study inclusion and exclusion criteria.

| **Table 1 Study inclusion and exclusion criteria** |
| --- |
| **Inclusion criteria** |
| · Age >12 years<br>**And** at least one of the following:<br>· Rhinorrhea<br>· Sore throat<br>· Sneezing (new)<br>· Cough (acute, chronic or productive)<br>· Wheeze<br>· Fever<br>· Shortness of breath<br>· New-onset hoarse voice |
| **Exclusion criteria** |
| · Lack of consent<br>· Ventilatory support<br>· Terminal disease<br>· Medical contraindication to voluntary coughing:<br>– Severe respiratory distress<br>– History of pneumothorax<br>– Eye chest or abdominal surgery in the past three months<br>· Uncontrolled acute heart failure (chronic heart failure was not an exclusion)<br>· History of neuromuscular disease<br>· History of lobectomy/pneumonectomy<br>· Diagnosed COPD or restrictive lung disease |

### Study protocol

The study did not interfere with clinical care. There were no adverse events reported.

#### Cough recording

Different operators collected cough recordings at the time of enrolment using iPhone 6 smartphones held 25-50cm from the mouth out of direct air flow. Recordings occurred in typical environments with background noises (talking, medical devices, footsteps and doors). We avoided recording coughs from other people or television sounds.

#### Clinical data

Data collected included the treating physician’s final diagnosis, demographics, medical history, presenting symptoms, vital signs, examination findings, response to treatments and results of investigations performed.

#### Clinical Examination

A full medical assessment was performed on all subjects at time of enrolment.

#### Clinical diagnosis of CAP (Reference Test)

Table 2 shows the definition of CAP (as per ERS) (15). The medical record for each patient was reviewed by an independent physician who took into consideration all available laboratory/radiology results and clinical examination findings to confirm a final clinical diagnosis. Pneumonia severity was assessed using the CRB-65 (19). After a clinical diagnosis was assigned the database was locked to further input.

**Table 2.** Clinical Diagnosis Definition.

|  |  |
| --- | --- |
| Community-Acquired<br>Pneumonia (CAP) | Must have both:<br>- New respiratory symptoms (SOB, cough, chest pain)<br>- Acute fever within the last week<br>- Treating team diagnosis of CAP and treated with antibiotics<br>Includes<br>· <u>Definite CAP</u>: Radiologist-reported new consolidation or significant infiltrate on the CXR or consolidation on CT.<br>· <u>Suspected CAP</u>: Either no CXR performed or no consolidation/infiltrate on CXR. |

#### Development of the algorithm (Index Test)

Between Jan 2016 and Nov 2017, we recruited subjects to train and refine the algorithm. The mathematical techniques used to derive our algorithms have been described elsewhere (16-18, 20). Briefly, selected cough audio-segments and patient-reported symptoms were extracted from the training cohort and combined into several continuous classifier models to determine the probability of CAP. A logistic-regression model was trained to diagnose CAP. The optimal model and corresponding probability decision thresholds were selected using a Receiver Operating Characteristic (ROC) curve with due consideration given to achieving a balance of PPA and NPA (20). The algorithm represents the weighted combination of clinical and cough derived features used.

#### Diagnostic accuracy study of the optimised diagnostic algorithm

We recruited subjects from the same locations as the training set, using the same inclusion/exclusion criteria, for the prospective diagnostic accuracy trial.

We used automatic segmentation to extract five coughs for analysis. For each subject, the algorithm reached a decision using audio-data plus input from four patient-reported symptoms: presence of fever in the past week, presence of either acute cough or productive cough and age. An independent operator ran the algorithm on the testing set to ensure blinding.

#### Statistical Analysis

As a clinical diagnosis of CAP is a not a gold-standard reference, we used PPA and NPA as primary measures of diagnostic agreement, rather than sensitivity or specificity. PPA reports clinical diagnosis-positive cases who are also index test positive; NPA reports clinical diagnosis-negative cases who are also index test negative. Results which were unsure (reference test) or missing (index test) were excluded from analysis.

Power calculations were derived based on expected PPA and NPA greater than 85% from the training program. A minimum of 48 cases were required. 95% confidence intervals were calculated using the method of Clopper-Pearson. The probability of a positive clinical diagnosis was calculated by the final classifier model and was used as the decision thresholds in derived ROC curves with AUC calculations.

Results are shown for the entire cohort and the following age groups: 12-22yrs.; >22yrs; 22- 65yrs and >65yrs. These age groupings are consistent with both FDA regulatory and CRB-65 guidelines (19, 21).

## RESULTS

### Diagnostic accuracy study

Between December 2017 and March 2019, 331 subjects were enrolled and completed the index and reference tests. Nine subjects were excluded because of recording issues or uncertain clinical diagnoses, leaving 322 subjects: 159 with CAP and 163 with non-pneumonic respiratory disease (figure 1). Of these, 200 came from the emergency department or inpatient wards and 122 from ambulatory care.

**Figure 1:**
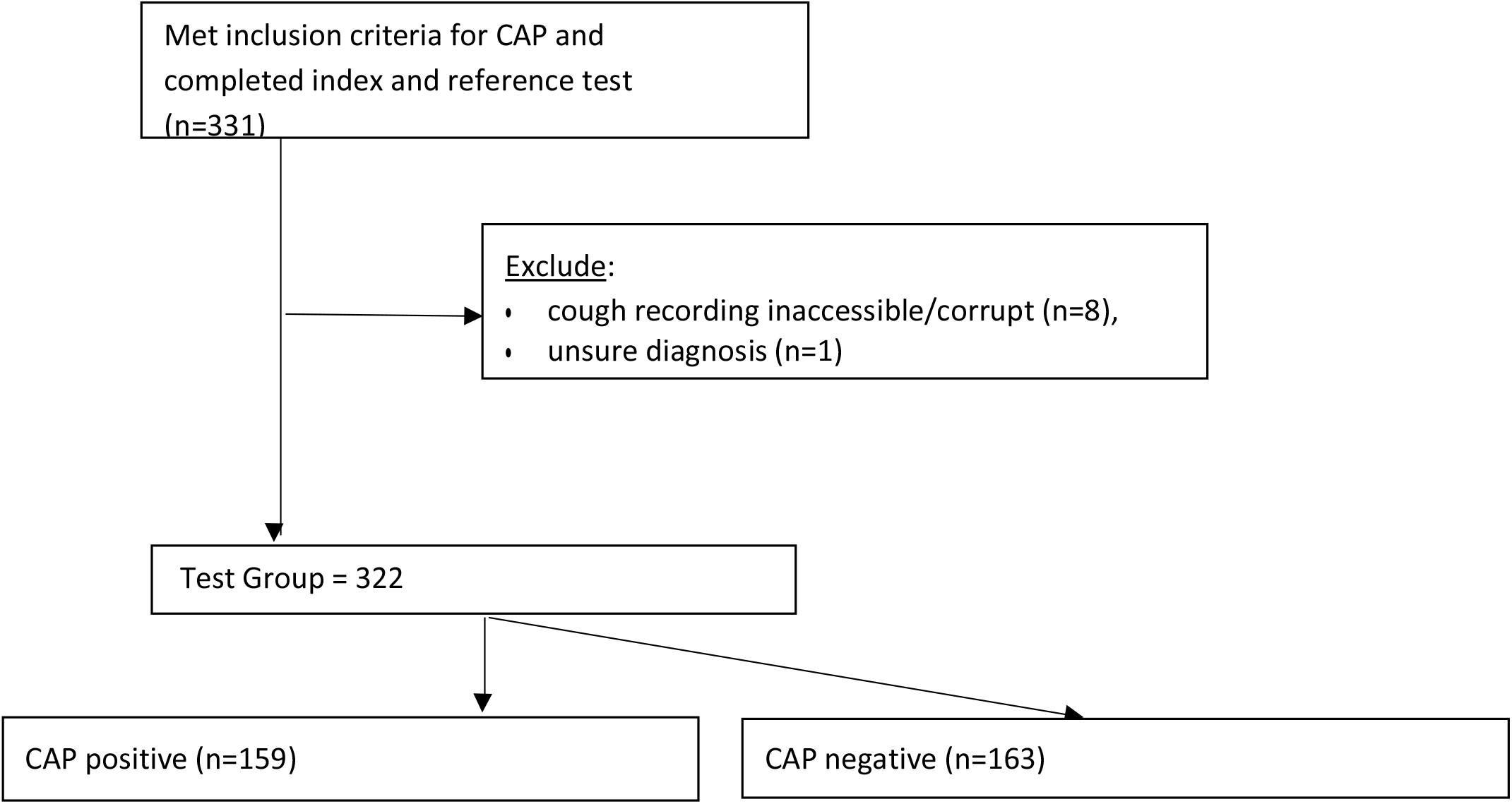
The flow of participants through the diagnostic accuracy study. Index test = software algorithm, reference test = clinician diagnosis.

Demographics and clinical features are shown in Tables 3 and 4. The mean age of all participants was 48.5±22.0yrs, 61.5% were female. There were more females than males for subjects >22yrs (62.6% vs 37.4%, p<0.01). There were no differences in past medical history between the 22-65yr and >65yr groups for chronic respiratory disease (p=0.279) or smoking history (p=0.974). Subjects > 65yrs were more likely to have comorbid chronic heart failure (p<0.009) or atopic history (p<0.001).

**Table 3.** Characteristics of participants included and excluded from the analysis. Data includes all subjects in each age group (CAP positive and negative)

|  | Total cohort<br>all ages<br>(n=322) | Age > 12 to<br><22 yr.<br>(n=44) | Age ≥ 22 yr.<br>(n=278) | Age ≥ 22 to<br><65 yr.<br>(n=192) | Age ≥ 65<br>yr.<br>(n=86) | Non-Analyzed.<br>(n=9) |
| --- | --- | --- | --- | --- | --- | --- |
| **Age (yr.)** |  |  |  |  |  |  |
| Mean ± SD | 48.5 ± 22.0 | 16.3 ± 3.1 | 53.6 ± 19.2 | 43.1 ± 12.4 | 77.0 ± 7.5 | 75.6 ± 16.1 |
| Median<br>(Q1, Q3) | 48.5 (30, 66) | 16 (13, 19) | 53(37, 69) | 43 (31, 54) | 76 (71, 84) | 79 (64, 89) |
| **Sex** |  |  |  |  |  |  |
| Male | 124 (38.5%) | 20 (45.5%) | 104 (37.4%) | 68 (35.4%) | 36 (41.9%) | 6 (66.7%) |
| Female | 198 (61.5%) | 24 (54.6%) | 174 (62.6%) | 124 (64.6%) | 50 (58.1%) | 3 (33.3%) |

**Table 4:** Symptoms and past medical history of participants. Data includes all subjects in each age group (CAP positive and negative)

|  | Total<br>cohort (all<br>age)<br>(n=322) | Age > 12<br>years to <22<br>years (n=44) | Age ≥ 22<br>years<br>(n=278) | Age ≥ 22<br>years to<br><65 years<br>(n=192) | Age ≥ 65<br>years<br>(n=86) |
| --- | --- | --- | --- | --- | --- |
| **Symptoms during this illness** |  |  |  |  |  |
| Acute Cough | 171 (53.1%) | 12 (27.3%) | 159 (57.2%) | 97 (50.5%) | 62 (72.1%) |
| Chronic Cough | 18 (5.6%) | 2 (4.6%) | 16 (5.8%) | 9 (4.7%) | 7 (8.1%) |
| Productive Cough | 114 (35.4%) | 9 (20.5%) | 105 (37.8%) | 61 (31.8%) | 44 (51.2%) |
| Fever | 145 (45.0%) | 13 (29.6%) | 132 (47.5%) | 83 (43.2%) | 49 (57.0%) |
| Runny Nose | 127 (39.4%) | 9 (20.5%) | 118 (42.5%) | 78 (40.6%) | 40 (46.5%) |
| SOB | 133 (41.3%) | 7 (15.9%) | 126 (45.3%) | 74 (38.5%) | 52 (60.5%) |
| Wheeze | 37 (11.5%) | 3 (6.8%) | 34 (12.2%) | 17 (8.9%) | 17 (19.8%) |
| Nausea/Vomiting | 65 (20.2%) | 9 (20.5%) | 56 (20.1%) | 40 (20.8%) | 16 (18.6%) |
| Diarrhoea | 30 (9.4%) | 4 (9.1%) | 26 (9.4%) | 18 (9.4%) | 8 (9.3%) |
| Lethargy | 158 (49.1%) | 12 (27.3%) | 146 (52.5%) | 89 (46.4%) | 57 (66.3%) |
| Loss of Appetite | 82 (25.5%) | 8 (18.2%) | 74 (26.6%) | 37 (19.3%) | 37 (43.0%) |
| Hoarse Voice | 55 (17.5%) | 5 (11.4%) | 50 (18.0%) | 34 (17.7%) | 16 (18.6%) |
| **Past medical history** |  |  |  |  |  |
| Chronic<br>Respiratory<br>Disease | 59 (18.3%) | 11 (25.0%) | 48 (17.3%) | 30 (15.6%) | 18 (20.9%) |
| Atopy | 51 (15.8%) | 4 (9.1%) | 47 (16.9%) | 40 (20.8%) | 7 (8.1%) |
| Heart Failure<br>(controlled) | 30 (9.3%) | 0 (0.0%) | 30 (10.8%) | 2 (1.0%) | 28 (32.6%) |
| Smoker (ever) | 131 (40.7%) | 9 (20.5%) | 122 (43.9%) | 84 (43.8%) | 38 (44.2%) |

**Table 5:** PPA and NPA and calculated AUC of the software algorithm compared with a clinical diagnosis of CAP.

|  | PPA (%)<br>[95% CI]<br>(n= CAP Positive) | NPA (%)<br>[95% CI]<br>(n= CAP negative) | AUC<br>[95% CI] |
| --- | --- | --- | --- |
| **CAP: Total cohort (n=322)** |  |  |  |
|  | 86.2% [79.8%, 91.1%]<br>(n=159) | 86.5% [80.3%, 91.3%]<br>(n=163) | 0.95 [0.93, 0.97] |
| **CAP: 22-65 YEARS (n=192)** |  |  |  |
|  | 85.7% [76.4%, 92.4%]<br>(n=84) | 87.00% [79.2%, 92.7%]<br>(n=108) | 0.94 [0.91, 0.97] |
| **CAP: ≥65 YEARS (n=86)** |  |  |  |
|  | 85.7% [75.3%,92.9%]<br>(n=70) | 87.5% [61.7%, 98.4%]<br>(n=16) | 0.95 [0.91, 0.97] |

## Discussion

### Summary

We have developed and tested a mathematical algorithm for diagnosing CAP using cough sound analysis and patient-reported symptoms. The algorithm showed a high degree of agreement with clinician diagnosis (utilising all clinical and imaging modalities) with accuracy maintained across age groups and severity indices. The tool does not rely on the input of vital signs, clinical examination or radiological findings; and delivers an immediate, point-of-care result.

### Strengths and limitations

The study design ensured the independence of the training/testing sets, and the clinical/algorithm diagnostic teams. We applied stringent criteria, utilising all available medical and imaging data, to determine a clinical diagnosis that was as accurate as possible. The reference diagnosis of CAP was confirmed after patient discharge, whereas the algorithm provided diagnosis at the time of presentation. We implemented this rigorous process as studies have shown up to 30% discordance between initial and final diagnoses of pneumonia (22-24).

This study builds on our work diagnosing pneumonia in children aged 1 month-12 years (PPA 85%, NPA 80%) (16). The numbers in this study exceeded that required by power calculations except in the 12-22yrs group. Although there was good agreement between the algorithm and clinical diagnoses, and we have no reason to consider that diagnostic accuracy would not be maintained across this age group, it would be helpful to confirm this in a larger study.

This study population was from a first-world metropolitan hospital. It will be important to replicate the study in general practice, digital and other settings to assess diagnostic accuracy and user experience. As the algorithm is site- and operator-independent, we believe the results of the study are generalisable to community use. It was encouraging to note the high rate of usable cough data-sets.

A diagnostic concern in elderly patients presenting with respiratory symptoms is differentiating acute heart failure from CAP. We were pleased to see that the accuracy was preserved in the >65yr group, which had a higher prevalence of chronic heart failure.

Judging the severity of CAP is crucial when managing CAP and can be determined using the CRB-65 as recommended in UK guidelines (14). CRB-65 uses an assessment of mental state and vital signs (including blood pressure) to stratify the severity of pneumonia and 30-day mortality in hospitals and suggest where management should occur (19). Using the CRB-65, 41% of our CAP-positive cohort scored 0 (1.2% risk of 30-day mortality, suitable for community treatment) and 59% scored 1 or 2 (8.1% risk of 30-day mortality, hospital referral recommended) (25). We found no differences in the diagnostic accuracy of the algorithm between these groups in our study. As we excluded patients with severe respiratory distress, the severity of CAP in this study represents the population seen in general practice despite being hospital sourced. We anticipate that it would be more difficult to clinically diagnose pneumonias of lower severity. The ability of our algorithm to identify lower severity CAP is encouraging for its potential use in primary care.

### Comparison with existing literature

Many authors have reported on the difficulties of diagnosing CAP, and this is reflected in inconsistent management guidelines. Most have focussed on the relative merits of clinical features, examination findings and CXR; with very few reporting on digital diagnostic tools (26). Our prior work has shown that it is possible to use identify cough-sound streams that are characteristic of different respiratory disorders, including lower versus upper airway disease, COPD, COPD exacerbations and asthma exacerbations in adults; and focal pneumonia, croup and bronchiolitis in children (16).

In the absence of radiology, general practitioners are reliant upon empirical diagnosis of CAP using auscultation or clinical signs resulting in a significant proportion (71%) of radiologically-confirmed CAP cases being missed in primary care (27). Clinical signs and symptoms have previously been shown to perform poorly in the diagnosis of CAP (6, 8). Our algorithm provides high diagnostic accuracy in the absence of radiology and without the need to examine the patient.

### Implications for practice

The NHS has advised it expects all patients to have the right to be offered digital-first primary care by 2023-24 with estimates that 150m face-to-face GP visits could be replaced by telehealth each year (28). In April 2020, during the COVID-19 pandemic, 71% of all GP visits in the UK were conducted remotely compared to 25% pre-pandemic. Up to half of telehealth consultations are for respiratory disease (29, 30).

During the pandemic, NICE recommended that clinicians not consult face-to-face or perform auscultation for CAP diagnosis unless essential. Instead, they suggest visual observation for breathlessness and cyanosis and using clinical “gestalt” or vital signs (including temperature, heart and respiratory rates) to rule out CAP (31, 32). This would be challenging over a telephone or video call.

We have demonstrated that our algorithm can accurately identify CAP of varying severity, without clinical or radiological inputs. Its rapid output and smartphone platform, make it suited for both traditional and digital consultations where it can assist in CAP diagnosis using only cough sounds and four patient-reported symptoms.

The algorithm is incorporated into a platform to identify multiple respiratory disorders and may enhance the capabilities of community telehealth services, with implications for reducing the duration of illness minimising complications and promoting good antibiotic stewardship. The platform has regulatory approval for use in Europe and Australia. It is available on Australian telehealth platforms.

With current recommendations to use digital consultations during the pandemic, as well as the uptake of remote consultations globally, this tool presents an opportunity to use digital diagnostics to enhance telemedicine consultations.

## Data Availability

The underlying codes are the property of ResApp Health and are not available. The datasets supporting the conclusion of this article are available on reasonable request from PP and JB. The cough recordings are not available but will be uploaded as an educational tool in the future.

## DECLARATIONS AND ACKNOWLEDGEMENTS

### Ethical Approval

Ethical approval was obtained from the Human Research Ethics Committees of Princess Margaret Hospital (2015030EP), Joondalup Health Campus (1501), Curtin University (HRE2018-0016) and The University of Queensland (2015000395 / 2016.01.179). All participants, parents or guardians signed a consent to participate form.

### Reporting

This work has been reported in accordance with the Standards for Reporting Diagnostic Accuracy studies (STARD).

## Acknowledgments

The authors wish to thank the many patients and their families who have given their time and enthusiasm for this work.

---

**Figure S1a.**
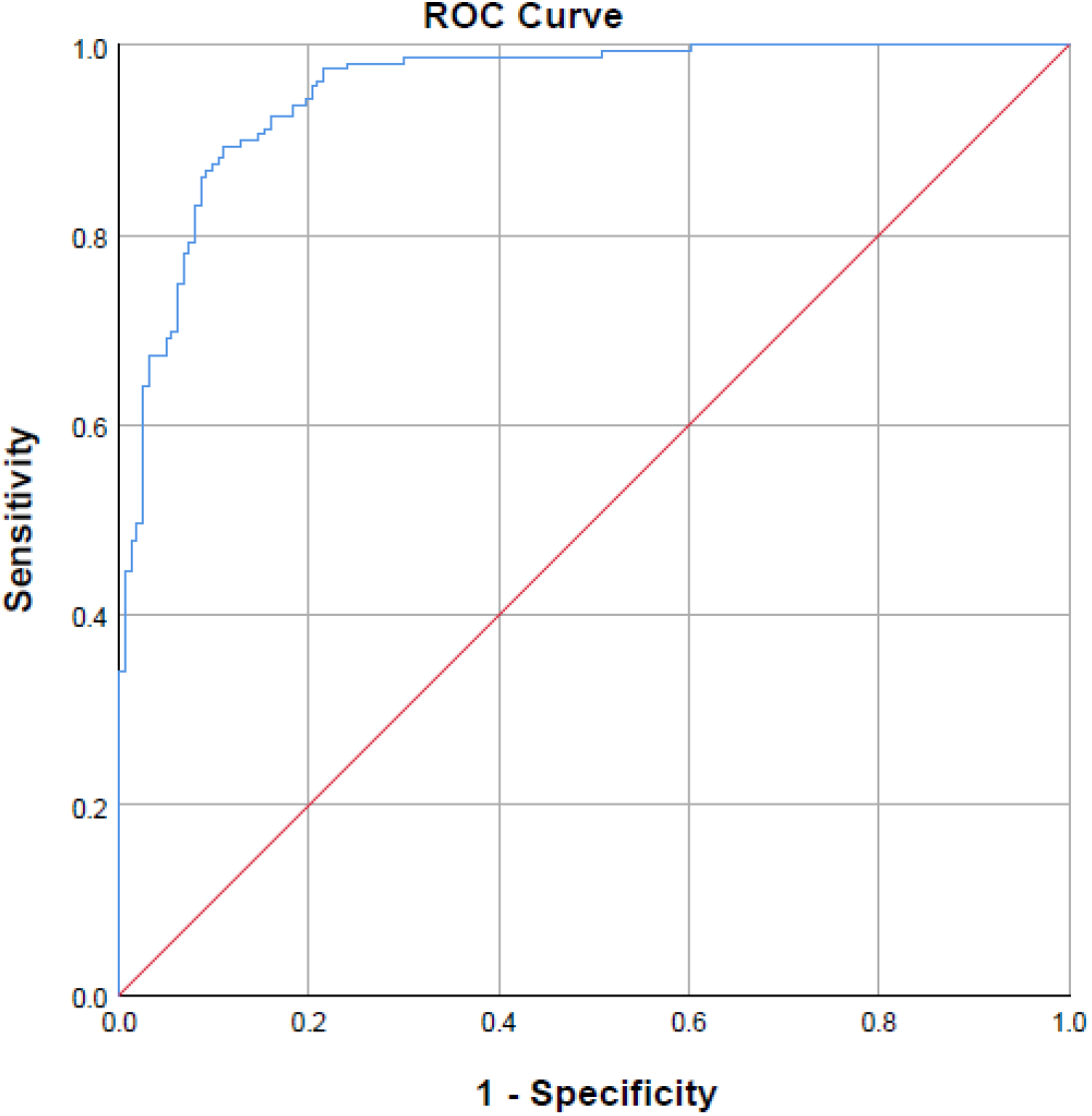
ROC for total cohort (n=322). AUC 0.95 [0.93,0.97].

**Figure S1b.**
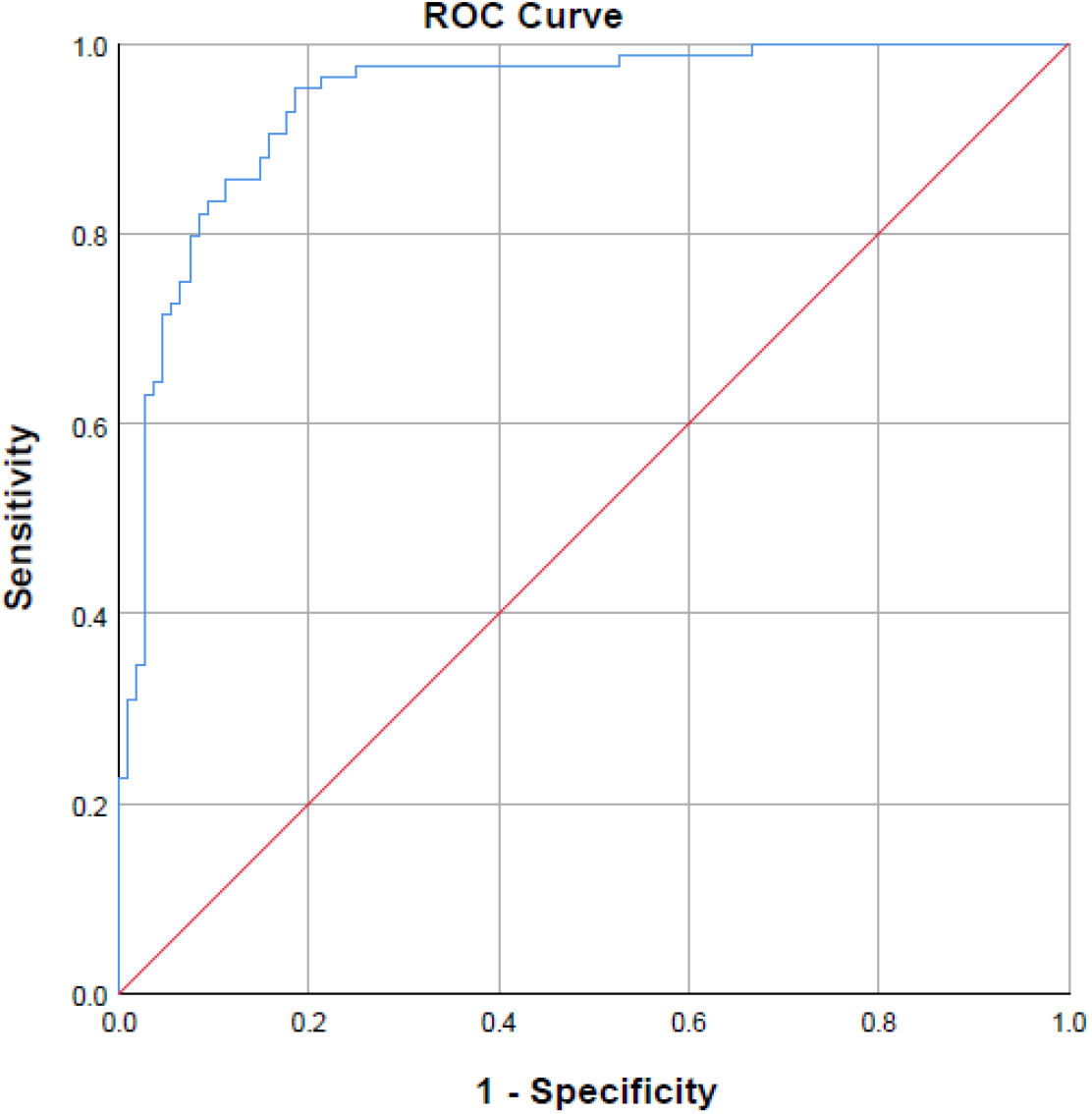
ROC for age 22-65 years (n=192). AUC 0.94 [0.91,0.97].

**Figure S1c.**
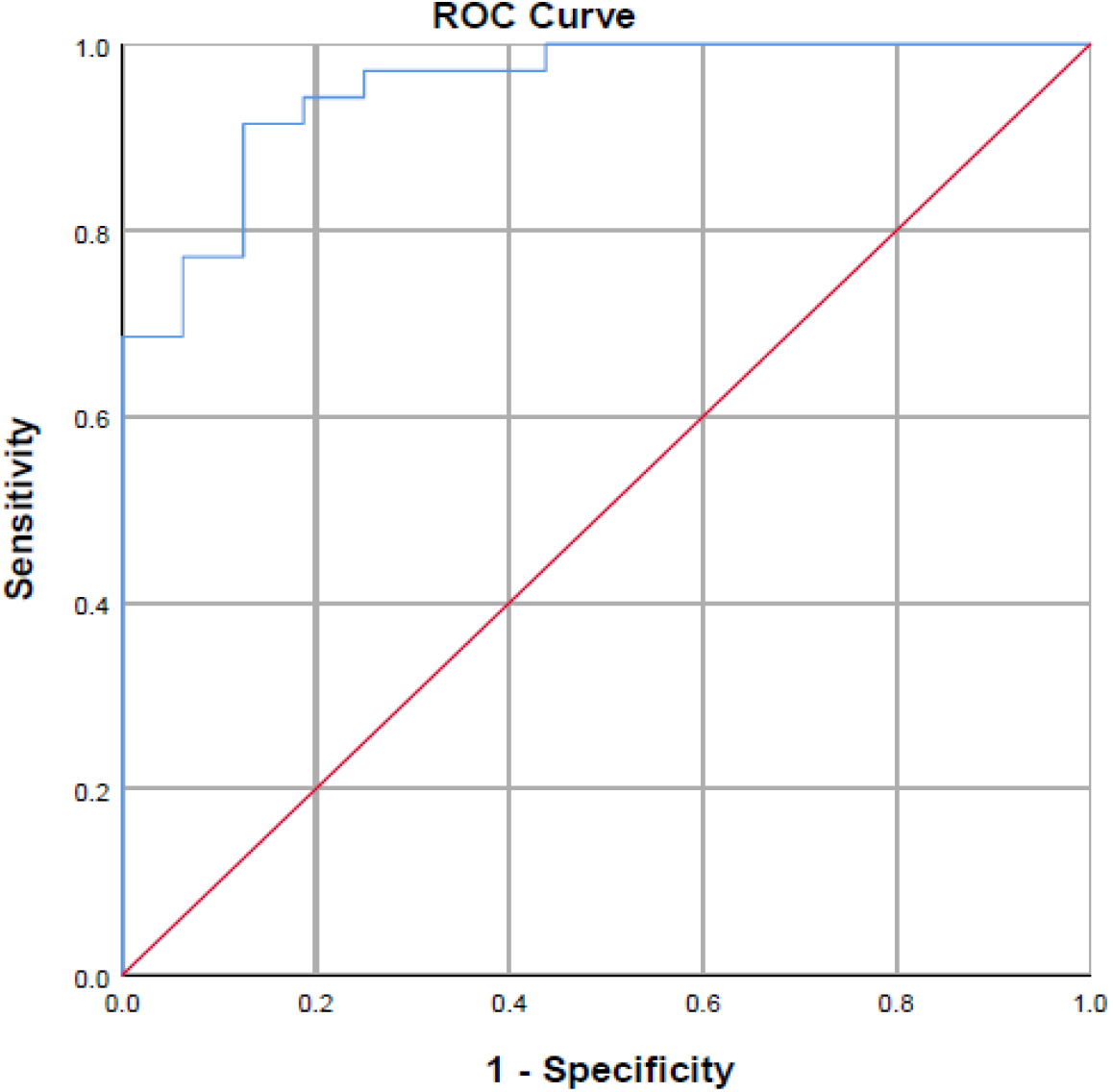
ROC for age > 65 years (n=86). AUC 0.95 [0.91,0.97].

